# Symptoms of Body Dysmorphic Disorder and Muscle Dysmorphia are Related to Greater Self-Face Recognition Difficulty and Negative Self-Face Evaluations Among Males

**DOI:** 10.1101/2025.01.29.25321224

**Authors:** Jade Portingale, Isabel Krug, David Butler

**Affiliations:** School of Psychological Sciences, The University of Melbourne, Melbourne, Australia; Faculty of Psychology and Counselling, The Cairnmillar Institute, Melbourne, Australia; Department of Psychology, Counselling and Therapy, LaTrobe University, Melbourne, Australia

**Keywords:** body dysmorphic disorder, muscle dysmorphia, male body image, face recognition, self-perception

## Abstract

**Objective:** Self-face perception is critical to physical self-concept, yet its importance in body image disorders among males is underexplored. This study examines how self-face recognition accuracy and evaluations are influenced by the severity of body dysmorphic disorder (BDD) and muscle dysmorphia (MD) symptoms in males and whether symptom severity moderates the relationship between recognition accuracy and evaluations

**Methods:** Sixty-eight White and East/Southeast Asian males in Australia completed measures assessing BDD and MD symptoms (appearance intolerance, drive for size, and functional impairment), self-face recognition accuracy (self-reported difficulty and objective sensitivity using a video-morphing task), and self-face evaluations (perceived attractiveness, adiposity, and dissatisfaction)

**Results:** Hierarchical regressions revealed that higher BDD symptoms and MD-related appearance intolerance predicted greater self-reported recognition difficulties and more negative evaluations (lower attractiveness, higher dissatisfaction). However, symptoms were unrelated to objective recognition sensitivity and perceived adiposity. Preliminary analyses suggested that recognition accuracy and evaluations were also unrelated, with no moderating effects of symptom severity.

**Conclusions:** These findings suggest that elevated BDD and MD symptoms, particularly appearance intolerance, disrupt self-face recognition and evaluations in males. Addressing these disturbances could enhance theoretical models of body image. Future research should investigate these processes in diverse clinical populations and longitudinal contexts before considering implications for intervention.

**Highlights:**

- Higher body dysmorphic disorder (BDD) symptoms were linked to greater self-face recognition difficulty and lower perceived facial attractiveness in males.
- Muscle dysmorphia (MD) symptoms, specifically appearance intolerance, related to greater self-face recognition difficulty, lower perceived facial attractiveness and greater head dissatisfaction.
- Preliminary evidence that self-face recognition difficulty and evaluations are unrelated, independent processes in males with BDD and MD symptoms.

## Introduction

Self-face perception broadly encompasses the recognition and evaluation of one’s face and is closely linked to self-consciousness, identity, and psychological well-being (e.g., Felisberti & Musholt, 2014). These processes may be inherently interlinked, with recognition accuracy influencing subsequent evaluations, while evaluative biases could affect basic recognition processes.

The significance of self-face perception has intensified in contemporary digital environments, where faces function as primary social identifiers through face-focused content, such as selfies (Fardouly & Rapee, 2019). The proliferation of facial editing tools and filters has created unprecedented opportunities to alter fundamental facial representations, potentially disrupting recognition and evaluation processes. Indeed, these technological advances have coincided with rising facial appearance concerns (Wang et al., 2021). However, research on self-face perception in body image disorders is critically lacking.

While a recent study highlighted impaired self-face recognition and negative evaluations in females with eating disorders (Portingale et al., 2024b), understanding of these processes in male-prevalent body image disorders remains limited. Given evidence that males experience distinct appearance pressures centred on facial masculinity (e.g., jaw definition, facial structure), investigation into self-face perception in male-prevalent disorders such as body dysmorphic disorder (BDD) and muscle dysmorphia (MD) is salient.

BDD is characterized by a persistent preoccupation with perceived appearance flaws that are negligible or unnoticeable to others (American Psychiatric Association [APA], 2024). Although this preoccupation in BDD often pertains to specific facial features (e.g., nose, eyes) (Phillips, 2005), relatively little attention has been given to how these populations process their own face holistically. Indeed, while current treatments address broader cognitive symptoms and distress, disturbances involving self-face perception remain critically unaddressed (Beilharz et al., 2017).

MD, a subtype of BDD predominately affecting males, is characterised by a pathological fixation on achieving muscularity and leanness (APA, 2024). Despite MD sharing many clinical features with BDD, literature on self-face perception in MD is entirely absent. Understanding how BDD and MD affect both the recognition and evaluation of one’s face could reveal important maintenance mechanisms and treatment targets.

### Self-Face Recognition in BDD and Implications for MD

A growing number of studies have implicated alterations in own-face processing in BDD (Beilharz et al., 2017), however, knowledge gaps remain. While some studies suggest that individuals with BDD (unlike controls) exhibit difficulties identifying unaltered photographs of their own faces (Buhlmann et al., 2014; Yaryura-Tobias et al., 2002), comprehensive investigations into broader self-face recognition accuracy (identifying one’s face as self) are lacking.

Several mechanisms potentially underlie these impairments. Impaired self-awareness is a common feature of BDD, associated with poor clinical insight and strong convictions about perceived flaws (Toh et al., 2017a), often coupled with alexithymia; a difficulty identifying and describing emotional states (Fenwick & Sullivan, 2011; Rosenberg et al., 2022). The connection between self-awareness and self-recognition (Suddendorf & Butler, 2013) suggests impaired self-awareness may hinder accurate self-face recognition.

Abnormal visual processing in BDD may further contribute to self-face recognition disturbances. While typical face recognition relies on holistic processing—integrating facial features into a unified whole (Van Belle et al., 2010)—BDD is characterized by feature-focused processing (Lang et al., 2021; Virgili et al., 2024). Eye-tracking studies reveal that individuals with BDD (unlike controls) display an attentional bias towards perceived flaws or unattractive features in their own faces (Greenberg et al., 2014; Grocholewski et al., 2012).

Reduced face inversion effects in BDD, a marker of impaired holistic processing, further support this pattern (Feusner et al., 2010; Toh et al., 2017c). Community samples with higher dysmorphic concern have also shown visual processing variations, including reduced face inversion effects (Beilharz et al., 2017), highlighting how impairments in self-face recognition may be observed across the continuum of BDD symptom severity.

Appearance-related compulsive behaviors in BDD and MD, such as mirror-checking and camouflaging (Buhlmann & Winter, 2017; Oakes et al., 2017), may exacerbate feature-focused processing and impair self-face recognition. Heightened anxiety during self-face viewing in BDD (Bohon et al., 2012) could further impair recognition, as anxiety is negatively associated with self-face recognition accuracy in the general population (Hills et al., 2016; Kim et al., 2016). Collectively, these mechanisms provide a basis for hypothesizing self-face recognition impairments in BDD, however, male-specific insights are lacking.

The male-specific research gaps in this domain are particularly concerning, given that MD predominantly affects males and involves unique visual processing biases related to muscularity ideals. Nonetheless, one eye-tracking study indicated parallels with BDD’s visual processing biases, documenting attentional biases toward perceived unattractive body areas in males with MD (Waldorf et al., 2019). MD and BDD also share compulsive behaviors like mirror-checking and camouflaging also observed in MD (Buhlmann & Winter, 2017). However, despite such potential mechanisms, self-face recognition in MD remains unexplored. Investigating self-face recognition is critical to understanding broader perceptual disturbances in BDD and MD, including self-face evaluations and relationships between the two processes.

### Self-Face Evaluations in BDD and Implications for MD

Maladaptive beliefs and negative emotions regarding one’s facial appearance are well-documented in BDD. Individuals with BDD typically overestimate others’ attractiveness while underestimating their own (Buhlmann et al., 2008; Lambrou et al., 2011). Greater dissatisfaction with facial appearance is consistently reported in individuals with BDD compared to those with eating disorders (Hrabosky et al., 2009; Toh et al., 2020).

Associations between facial dissatisfaction and dysmorphic concerns are also observed in community samples (Ayase et al., 2023). However, most studies have focused on females, limiting male-specific insights.

Body size estimation, including face size, is an especially novel area that remains unexamined in BDD and MD (Möllmann et al., 2024a). In males, negative self-face evaluations may stem from a complex interplay between muscularity ideals and facial morphology. Facial adiposity, a visible marker of body composition, significantly influences perceived attractiveness (de Jager et al., 2018; Lei et al., 2019). Elevated perceptions of facial adiposity in BDD may result from a tendency to overestimate flaws, while in MD, concerns about leanness may exacerbate such perceptions (APA, 2024; Buhlmann et al., 2008). These distorted perceptions may contribute to dissatisfaction with facial attractiveness and satisfaction.

Furthermore, the relationship between self-face recognition and evaluations remains underexplored in body image disorders. Evidence from one female-focused study suggests that recognition difficulties correlate with negative self-evaluations (Portingale et al., 2024b). However, whether these relationships manifest similarly in males with BDD or MD remains unknown. Elevated symptom severity may amplify the link between recognition deficits and negative evaluations, warranting further investigation into these mechanisms.

### The Current Study

This study aims to address three key gaps in the literature by examining the relationship between BDD and MD symptom severity with self-face recognition accuracy and self-face evaluations, and the potential moderating effects of symptom severity on the connection between recognition and evaluation. It was hypothesized that higher BDD or MD symptoms would be: (i) associated with poorer self-face recognition accuracy (H1); (ii) associated with more negative self-face evaluations (e.g., higher perceived adiposity, lower attractiveness, greater dissatisfaction) (H2); and (iii) predictive of a stronger relationship between recognition deficits and negative evaluations, moderated by symptom severity (H3).

Given the multidimensional nature of MD symptoms, this study separately assessed cognitive-affective appearance intolerance, behavioral drive for size, and functional lifestyle impairment (Hildebrandt et al., 2004) to explore their unique influences on self-face perception. By investigating a continuum of symptoms, the study aims to elucidate how varying levels of BDD and MD symptoms in males affect self-face recognition and evaluations, potentially informing targeted interventions that address perceptual and evaluative disturbances.

## Method

### Participants

The Australia-based community sample comprised 68 males, recruited through university participant pools, noticeboards, social media, snowballing, and personal contacts. Eligibility criteria included being at least 18 years old, proficient in English, identifying as male, and self-reporting White or East/Southeast Asian ethnicity, reflecting the ethnic congruency needed for stimulus models (Tajadura-Jiménez et al., 2012). Participants were predominantly young, with a BMI within the healthy range, and mostly identified as White, heterosexual, single, and students who had typically completed a Bachelor’s degree. Ethical approval was granted by a Melbourne-based university (ID: 2056250.1).

### Measures

#### Demographic

Participants self-reported age, height, weight (to calculate BMI), ethnicity, primary language, sexual orientation, marital status, employment status, and history of BDD diagnosis.

#### Body Dysmorphic Disorder Symptomatology

The Dysmorphic Concern Questionnaire (DCQ) assessed BDD symptoms using 7 items (e.g., “*Have you ever been very concerned about some aspect of your physical appearance?*”), rated on a 4-point scale (0 = *not at all*, 3 = *much more than most people*) (Oosthuizen et al., 1998). Higher total scores (range 0–21) indicate greater concerns. A score ≥ 9 identifies individuals at risk of BDD (Mancuso et al., 2010). Internal consistency in the present sample was good (α =.845).

#### Muscle Dysmorphia Symptomatology

The Muscle Dysmorphic Disorder Inventory (MDDI) measured MD symptoms with 13 items (e.g., “*I think my body is too small”*) rated on a 5-point scale (1 = *never*, 5 = *always*) (Hildebrandt et al., 2004). Total scores (range 13–65) and subscale scores (drive for size [5 items, range 5–25], appearance intolerance [4 items, range 4–20], functional impairment [4 items, range 4–20]) reflect MD symptom severity. A cut-off score > 39 denotes MD risk (Zeeck et al., 2018). Internal consistency in the current sample was acceptable to good for the total and subscale scores (α =.728 to.866).

#### Self-Face Recognition Accuracy

The Self-Face Recognition Questionnaire (SFRQ; Larøi et al., 2007) assessed subjective difficulties in self-face recognition via 3 items (e.g.,

“*When looking at myself in a mirror, a window, a video, or on a photo, I have sometimes mistaken my face for someone else’s face”)*. Responses were binary (1 = *yes;* 0 = *no*), with affirmative answers followed by ratings for difficulty frequency (0 = *very rarely*; 3 = *very often*), stress (0 = *not at all*; 3 = *very*) and tiredness (0 = *not at all*; 3 = *very*). Higher total scores (range 0–30) indicate greater difficulties. Internal consistency in our sample was good (α =.826).

The Self-Face Recognition Task (SFRT) employed video-morphing to objectively measure self-face recognition sensitivity (Panagiotopoulo et al., 2017; Tsakiris, 2008).

Participants viewed a 100-frame (1 sec/frame) morphing sequence transitioning from an unfamiliar, ethnicity-matched male face to their own (i.e., from 0% Self to 100% Self) and stopped the video when the image appeared more like themselves. Higher percentages of “self” indicated reduced sensitivity. Standardized procedures ensured consistent stimulus preparation (for details see Supplementary Material).

#### Facial Attractiveness and Adiposity

Participants rated their perceived facial attractiveness and adiposity using two 7-point scales (0 = *very unattractive/underweight*; 6 = very *attractive/overweight*) (Coetzee et al., 2012; Tinlin et al., 2013). Higher scores reflected greater perceived attractiveness and adiposity.

#### Head Dissatisfaction

The head subscale of the Body Satisfaction Scale (BSS) assessed satisfaction with seven facial features (e.g., face, jaw, teeth) using a 7-point scale. Higher scores (range 7–49) indicated greater dissatisfaction. Internal consistency in the current study was good (α =.844).

#### Alexithymia Symptomatology

The 20-item Toronto Alexithymia Scale (TAS-20; Bagby et al., 1994) assessed alexithymia symptomatology (difficulty identifying feelings, difficulty communicating feelings, and externally-oriented thinking) on a 5*-*point scale (1 = *strongly disagree*; 5 = *strongly agree*). Higher total scores (range 20–100) indicate greater alexithymia symptoms (α =.797).

### Procedure

Participants attended a one-on-one virtual session conducted via Zoom. After providing online consent, they completed baseline questionnaires (DCQ, MDDI, SFRQ, TAS-20) and the SFRT. The number of seconds at which the movie was stopped was recorded by the researcher. Next, participants completed the remaining measures (facial attractiveness/adiposity scales, BSS) in a randomized sequence. Sessions lasted approximately 25 minutes, and participants received a $30 AUD e-gift card for their time.

### Data Analytic Plan

#### Design and Statistical Analyses

Analyses were conducted in R Studio (Version 4.4.1). Hierarchical multiple regressions tested the following hypotheses: (i) BDD/MD symptoms (DCQ and MDDI total and subscale scores) predicting self-recognition accuracy (SFRQ and SFRT); (ii) BDD/MD symptoms total and subscale scores predicting self-face evaluations (attractiveness, adiposity, dissatisfaction); and (iii) moderating effects of BDD/MD symptoms (total scores) on self-face recognition accuracy-evaluation relationships using interaction terms. Covariates (age, BMI, ethnicity) were included in Step 1 given their significance in the BDD/MD and self-perception literature (Bjornsson et al., 2013; Haider et al., 2023; Mellor et al., 2013), Alexithymia, a self-face perception and BDD-related psychological construct (Fenwick & Sullivan, 2011; Rosenberg et al., 2022), was added as a covariate in Step 2. Continuous measures of BDD and MD symptoms were entered in Step 3. For H2 and H3, self-recognition accuracy measures were added in Step 4, with interaction terms included in Step 5. Separate models were run for each hypothesis.

#### Sample Size Calculation

An *a-priori* power analysis using *G*\*Power (Faul et al., 2007) indicated that a minimum of 55 participants was required for 80% power to detect a medium effect size (*f* ^2^ =.15, Cohen, 2013) at α =.05 (two-tailed) in linear multiple regression with a maximum of six predictors. A medium effect size was chosen due to feasibility, as smaller effects would require an unachievable sample size (*N* = 395) in this under-researched field, where recruitment is challenging. Our sample (*N* = 68) exceeded this requirement, providing sufficient power for main effects. However, as the current study was underpowered for detecting interaction effects, the moderation analyses were considered preliminary and exploratory.

#### Pre-Registration

The study’s design, hypotheses and analysis plan were pre-registered on the Open Science Framework (https://doi.org/10.17605/OSF.IO/9NQB3). Analyses deviated slightly, as body-perception outcomes were excluded due to scope limitations.

## Results

### Data Cleaning and Preparation

The final dataset (*N* = 68) met most statistical assumptions, including linearity, homoscedasticity, multicollinearity (no correlations >.9), and residual normality. Potential homoscedasticity violations in models predicting facial adiposity with BDD and MD symptoms (Breusch-Pagan p <.05) were addressed with robust standard errors. Missing SFRT values (10 cases) were determined to be MCAR (MCAR test p =.798; Hawkins test p =.831) and handled via multiple imputation (m = 5) for direct effects analysis. Due to complexity, moderation analyses used complete cases (N = 58).

### Sample Characteristics and Descriptive Statistics

Tables 1 and 2 summarize the sample’s characteristics and descriptive statistics. No participants reported lifetime BDD or MD diagnoses, and average alexithymia scores were near the scale midpoint. While most participants scored below the midpoint on BDD and MD symptoms, 18% and 15% were classified as at risk for BDD and MD, respectively. Among MD subscales, appearance intolerance slightly exceeded the midpoint, while drive for size and functional impairment remained below it.

**Table 1.**
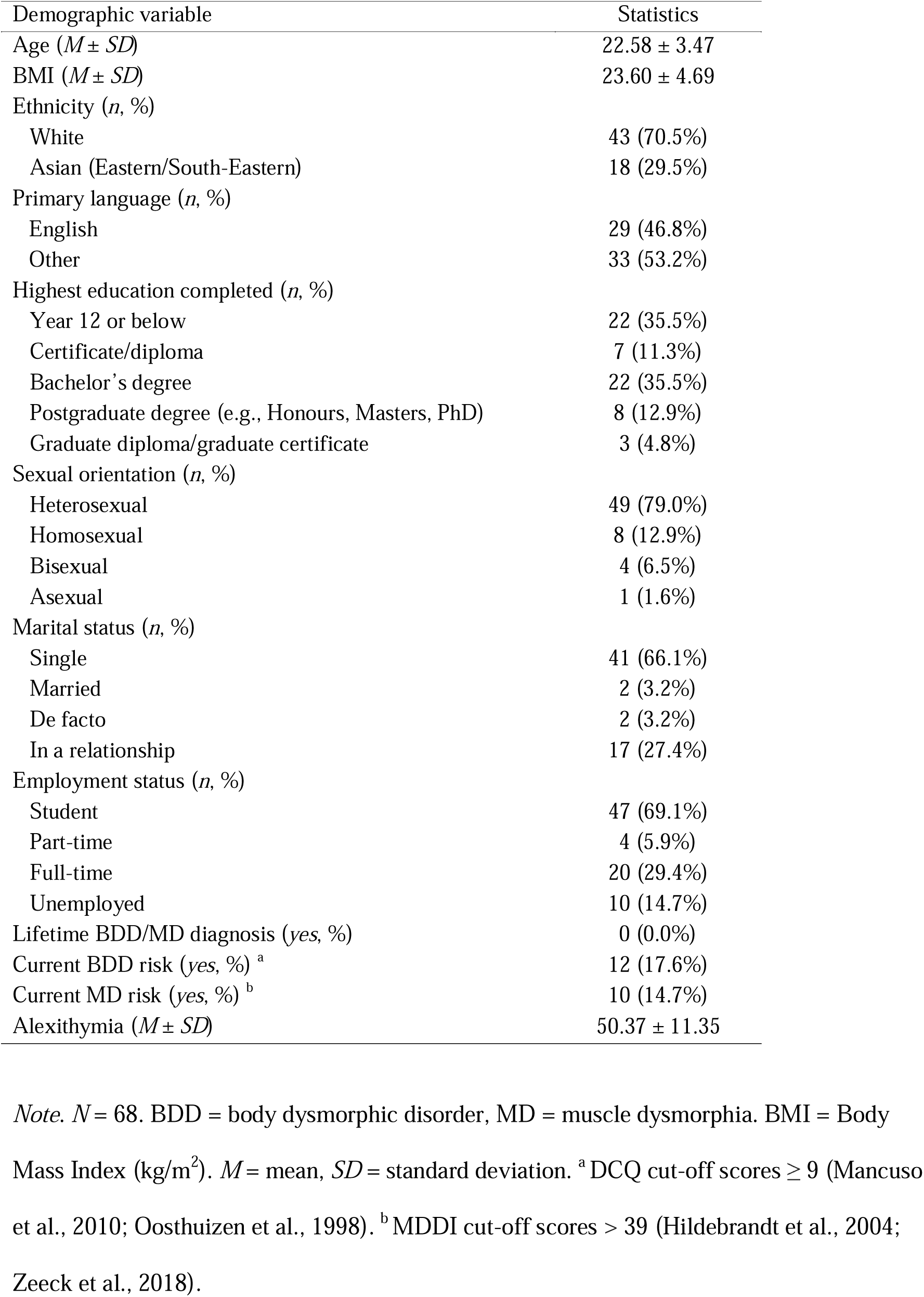
Demographic Characteristics.

**Table 2.** Descriptive Statistics for Modelled Variables.

| Variable | *M* | *SD* | Observed range | Possible range |
| --- | --- | --- | --- | --- |
| BDD symptoms <sup>a</sup> | 6.06 | 3.97 | 0–20 | 0–21 |
| MD symptoms (total score) <sup>b</sup> | 31.87 | 7.66 | 16–49 | 13–65 |
| Drive for size (subtype score) <sup>c</sup> | 13.16 | 4.80 | 5–24 | 5–25 |
| Appearance intolerance (subtype score) <sup>c</sup> | 10.51 | 3.83 | 4–19 | 4–20 |
| Functional impairment (subtype score) <sup>c</sup> | 8.19 | 3.66 | 4–20 | 4–20 |
| Self-face recognition accuracy |  |  |  |  |
| SFRQ (difficulty) | 3.75 | 3.89 | 0–18 | 0–30 |
| SFRT<sup>^</sup> (sensitivity) | 54.39 | 11.70 | 28.43–84.19 | 0.00–100.00 |
| Self-face evaluations |  |  |  |  |
| Facial attractiveness | 3.08 | 1.42 | 0–6 | 0–6 |
| Facial adiposity | 3.17 | 1.06 | 1–6 | 0–6 |
| Head dissatisfaction | 23.40 | 8.36 | 7–42 | 7–49 |

*Note.* *N* = 68. <sup>^</sup> % of self. BDD = body dysmorphic disorder; MD = muscle dysmorphia; SFRT = self-face recognition task; SFRQ = self-face recognition questionnaire. *M* = mean, *SD* = standard deviation. Higher scores on SFRT and SFRQ index poorer self-face recognition accuracy (i.e., poorer sensitivity and greater difficulty). <sup>a</sup> Dysmorphic Concern Questionnaire (DCQ; Oosthuizen et al., 1998) total score; <sup>b</sup> Muscle Dysmorphic Disorder Inventory (MDDI; Hildebrandt et al., 2004) total score; <sup>c</sup> MDDI subtype scores.

Participants required a higher percentage of their own face for accurate recognition (indicating reduced sensitivity) based on SFRT scores, while SFRQ scores suggested low difficulty in self-face recognition. Perceptions of facial attractiveness and adiposity were slightly above average, and head dissatisfaction scores were just below the midpoint.

### Hypothesis Testing

Table 3 presents results for H1–H3 using total symptom scores for BDD and MD, including self-face recognition and evaluation outcomes (3-step models), accuracy-evaluations relationship (4-step models), and moderation analyses (5-step models). Table 4 extends the analyses for H1 and H2 by incorporating subscale scores for MD symptoms (appearance intolerance, drive for size, and functional impairment).

**Table 3.**
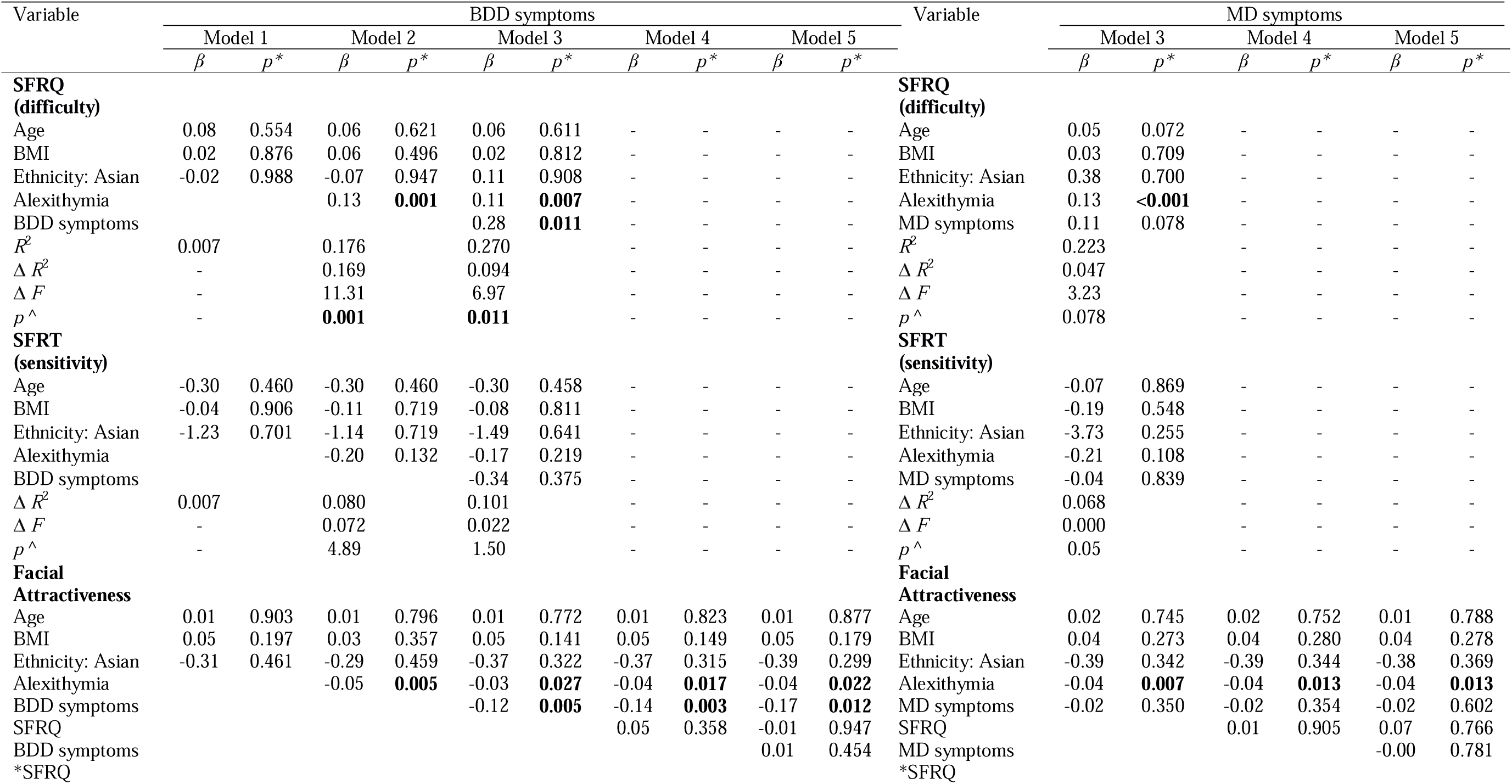
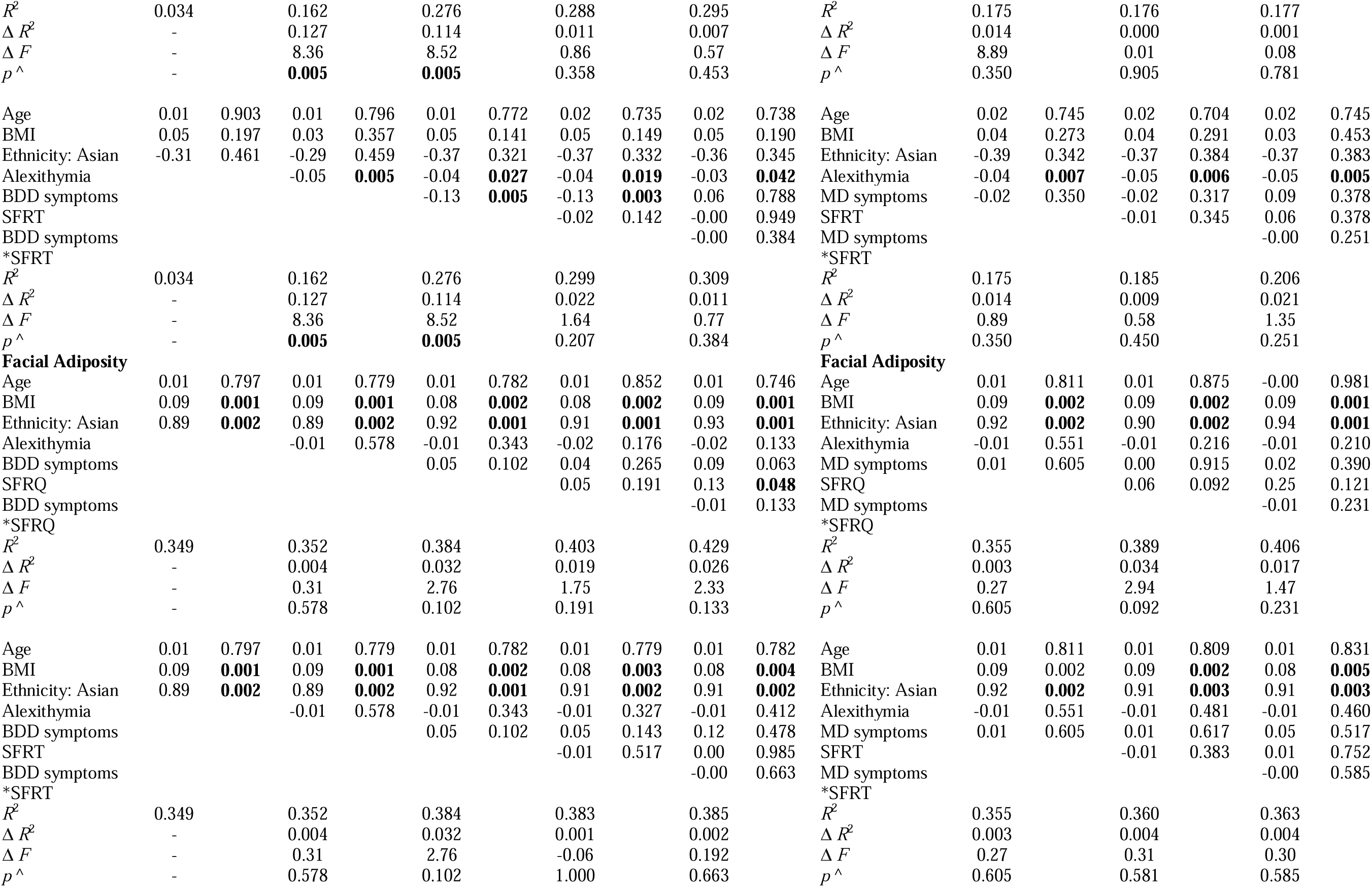
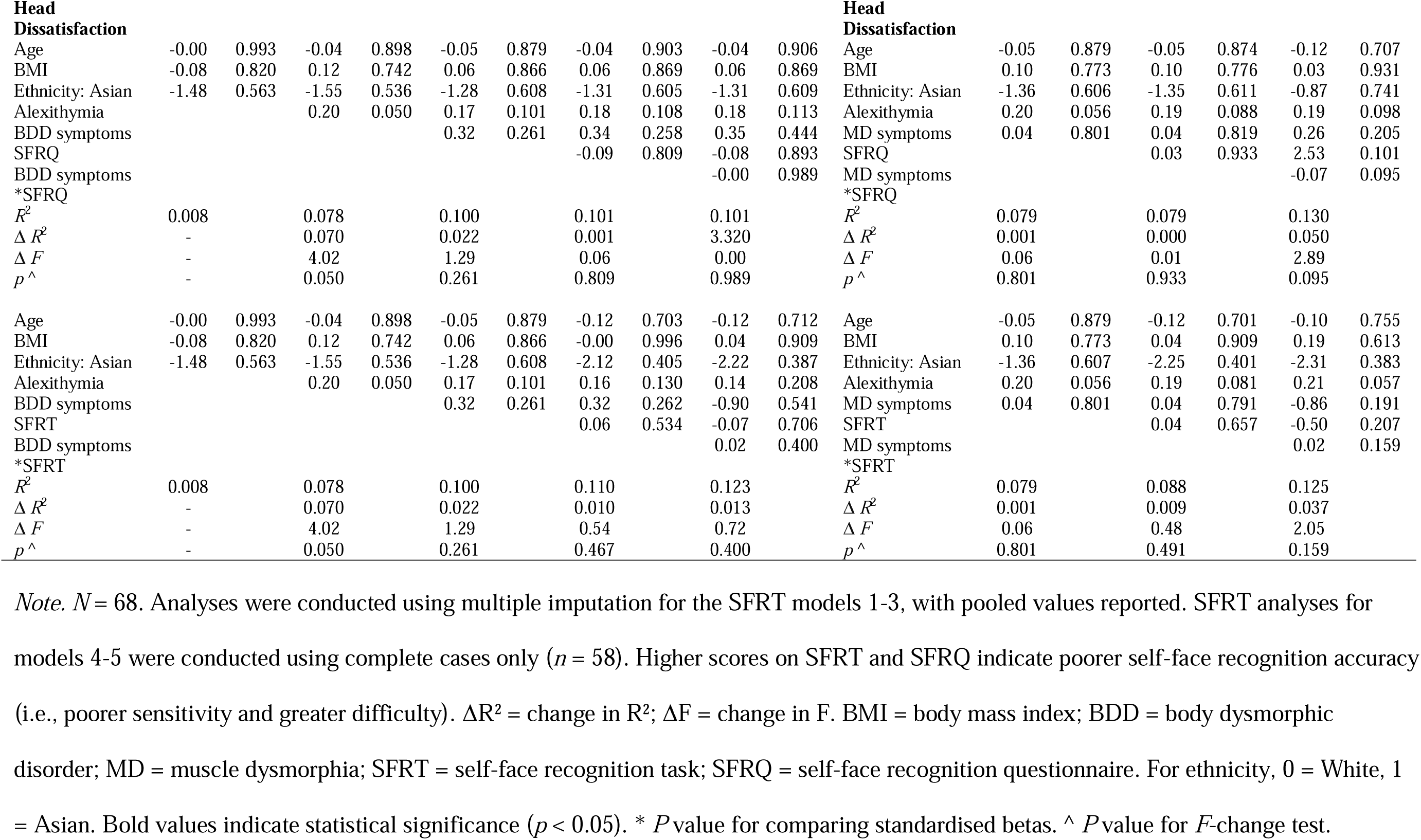
Modelled Relationships Between Overall BDD and MD Symptoms with Self-Face Recognition Accuracy (H1) and Self-Face Evaluations (H2), and Moderation Effects (H3).

**Table 4.**
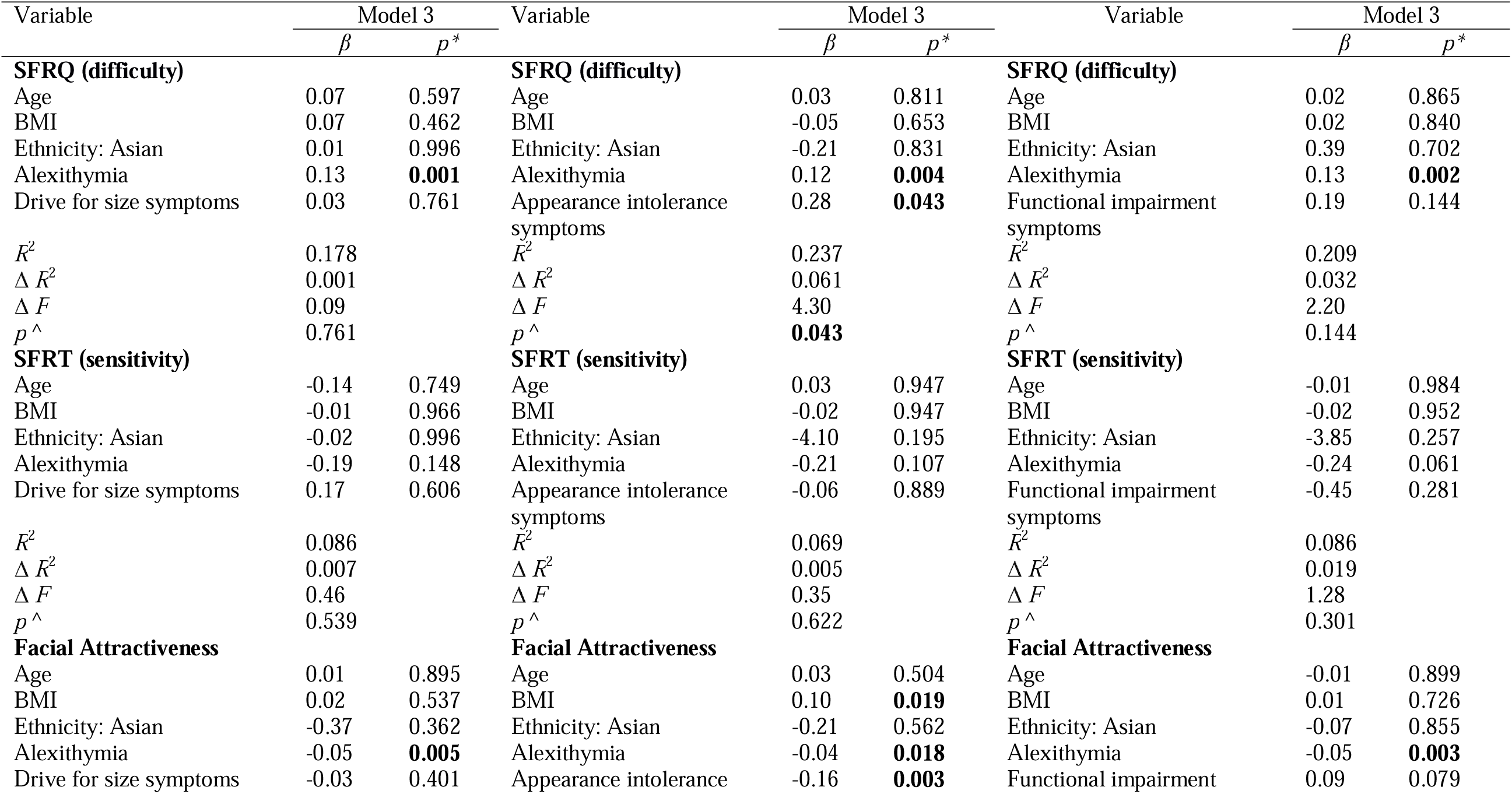
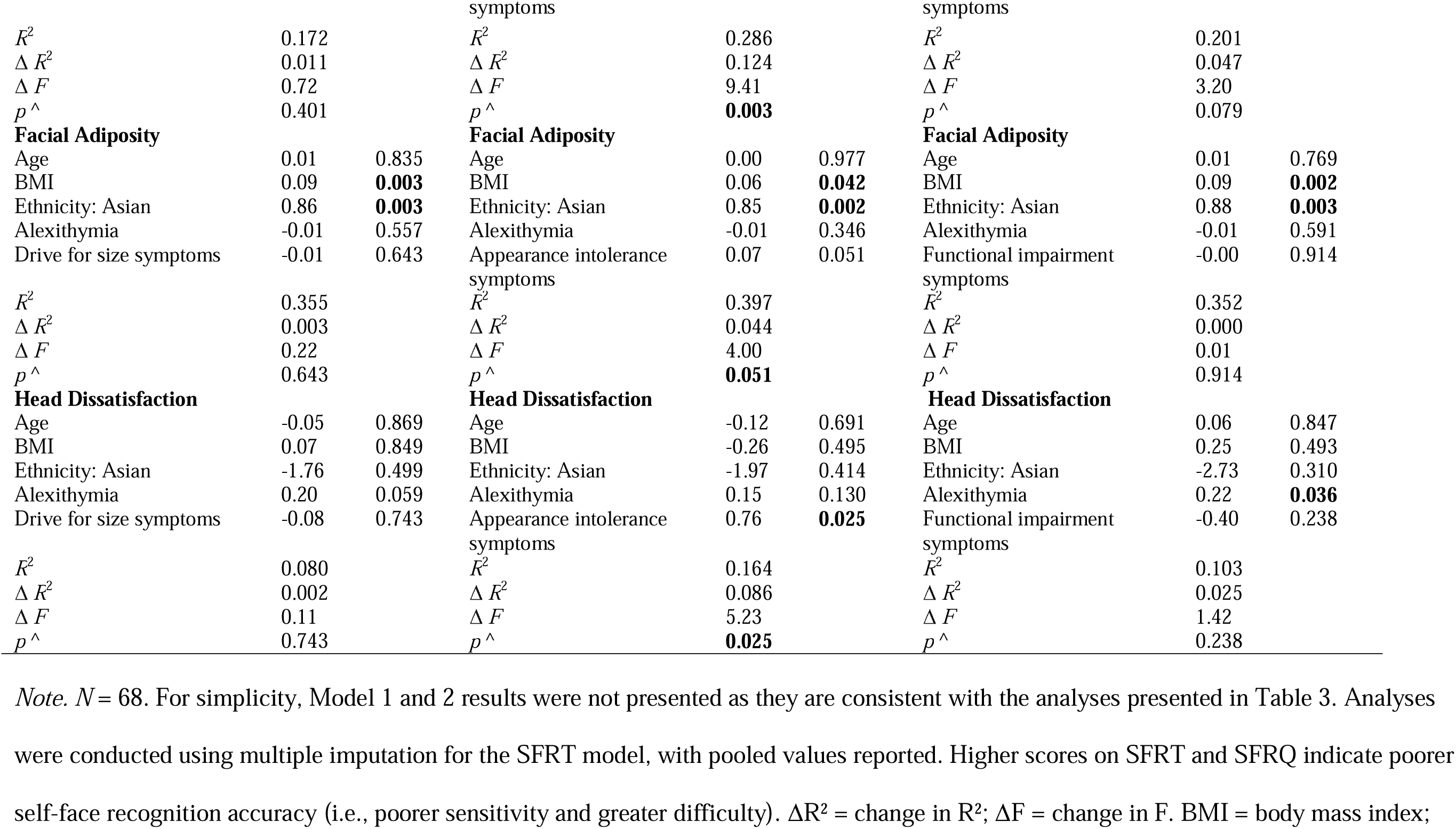
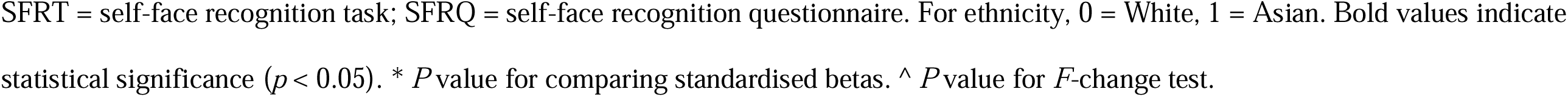
Modelled Relationships Between MD Symptom Subtypes (Drive for Size, Appearance Intolerance, Functional Impairement) with Self-Face Recognition Accuracy (H1) and Evaluations (H2) (Model 3)

#### H1: BDD and MD Symptoms and Self-Face Recognition Accuracy

Higher BDD symptoms significantly predicted greater self-face recognition difficulty (SFRQ; 9% unique variance, 27% total variance explained). For SFRT, BDD symptoms (3% unique variance) and covariates were non-significant (8% total variance explained).

Total MD symptoms were unrelated to SFRQ scores (22% total variance explained, 9% unique variance) or SFRT scores (6% total variance explained, 0% unique variance).

However, MD subscales showed that appearance intolerance was associated with greater SFRQ scores (6% unique variance, 24% total variance). Drive for size and functional impairment were unrelated to SFRQ scores (18-21% variance explained, 0-3% unique variance). MD subscale scores were unrelated to SFRT scores (6-9% variance explained, 0%-2% unique variance).

#### H2: BDD and MD Symptoms and Self-Face Evaluations

BDD symptoms were significantly associated with lower perceived facial attractiveness (11% unique variance, 27% total variance explained). BDD symptoms were not significantly associated with perceived facial adiposity or head dissatisfaction (38% and 10% variance explained, 3% and 2% unique variance, respectively).

Total MD symptoms were unrelated to evaluations (8-36% total variance explained, 0-1% unique variance). However, appearance intolerance was associated with lower perceived attractiveness (12% unique variance, 29% total variance) and higher head dissatisfaction (9% unique variance, 16% total variance). Drive for size and functional impairment subscales were non-significantly related to evaluations (8-36% variance explained, 0-4% unique variance).

#### H3: Self-Face Recognition Accuracy-Evaluations Relationship, Moderated by BDD and MD Symptoms

Preliminary analyses demonstrated that recognition accuracy (SFRQ or SFRT) was unrelated to evaluations, and BDD/MD symptoms did not moderate the accuracy-evaluation relationships (0-5% unique variance explained).

#### Covariates

In some models, higher levels of alexithymia were associated with greater recognition difficulty and lower perceived facial attractiveness. Higher BMI and Asian ethnicity were associated with higher perceived adiposity.

## Discussion

This study examined relationships between BDD and MD symptoms, self-face recognition accuracy, and self-face evaluations in males. The results indicate that higher BDD and domain-specific MD symptoms (i.e., appearance intolerance) were linked to greater difficulty in self-face recognition and more negative evaluations of facial attractiveness and head dissatisfaction. Contrary to expectations, however, BDD and MD symptoms did not affect sensitivity in detecting one’s own face or perceived facial adiposity, and accuracy-evaluation relationships were not moderated by symptom severity. However, as the latter moderation analyses were underpowered, these results were considered preliminary and interpreted with caution.

### Self-Face Recognition Accuracy in BDD and MD

Participants with higher BDD and MD symptoms exhibited unique patterns of self-face recognition difficulty and sensitivity. Specifically, self-face recognition difficulty was linked to heightened BDD symptoms, accounting for 9% unique variance. This aligns with previous research indicating that BDD involves impaired visual processing for personally relevant stimuli like one’s own face (Buhlmann et al., 2014; Yaryura-Tobias et al., 2002). This difficulty may stem from an overreliance on detailed-oriented processing at the expense of holistic integration, a characteristic previously observed in BDD populations (Lang et al., 2021; Virgili et al., 2024). Such processing biases could lead individuals to misidentify their own faces.

For MD, only appearance intolerance symptoms were linked to recognition difficulty, accounting for 6% of the variance. Unlike more behaviorally-and functionally-oriented MD symptoms, this symptom cluster—characterised by appearance anxiety and negative beliefs

(Hildebrandt et al., 2004)—may be more directly related to self-perception. This finding aligns with evidence that anxiety impairs self-face recognition accuracy in the general population (Hills et al., 2016; Kim et al., 2016) and highlights the importance of examining symptom clusters within MD in self-perception research. Additionally, attentional biases in MD toward unattractive body areas (Waldorf et al., 2019) may extend to facial features, impairing holistic processing required for accurate recognition, similar to BDD.

In contrast, self-face recognition sensitivity was unrelated to BDD or MD symptoms.

The dissociation between subjective difficulty and objective sensitivity offers insights into the cognitive mechanisms underlying body image disorders. The self-report questionnaire (SFRQ) taps into long-term self-representations, potentially influenced by distorted memory representations and emotional biases. Indeed, there is evidence of impaired long-term memory performance for recognising facial stimuli in individuals with self-reported BDD versus controls (Möllmann et al., 2024b). Meanwhile, the objective task (SFRT) relies more on immediate visual processing and may be less affected by biases in stored representations. Anxiety—shown to impair self-face recognition (Hills et al., 2016; Kim et al., 2016)—may mediate these effects, with the subjective measure potentially amplifying anxiety-driven preoccupation with appearance, whereas the time-constrained task may reduce anxiety-driven distortions. Nonetheless, we acknowledge that self-face viewing often exacerbates anxiety in BDD (Bohon et al., 2012). By informing participants they would view their own image, we may have inadvertently excluded individuals with more severe BDD symptoms who experience anxiety during self-face viewing, potentially biasing the sample.

The null findings for the SFRT may also be due to methodological limitations.

Hong Liu and Chen (2018) demonstrated how placing an oval frame around facial images used in a video-morphing task (consistent with our approach) impairs holistic processing. Hence, our study may have constrained all participants to engage in feature-based processing, potentially masking nuanced visual processing differences associated with BDD (Virgili et al., 2024).

### Self-Face Evaluations in BDD and MD

Higher BDD symptoms were associated with lower perceived facial attractiveness, explaining 11% of unique variance. This finding extends previous research on negative self-evaluations in BDD (e.g., Buhlmann et al., 2008; Lambrou et al., 2011) to a male-specific sample. For MD, appearance intolerance was the sole MD symptom linked to negative evaluations, explaining 9-12% of the variance. This novel finding expands the understanding of cognitive-affective disturbances in MD to facial appearance, supporting theoretical models that emphasize the importance of appearance intolerance in body image disturbances (Hildebrandt et al., 2004). Theoretically, the feature-based processing bias towards perceived unattractive facial features characteristic of BDD (Virgili et al., 2024) and potentially MD (Waldorf et al., 2019) may promote negative self-face evaluations by disrupting integrated perceptual experiences.

Unexpectedly, perceived facial adiposity was unrelated to BDD or MD symptoms.

This finding may reflect the specific nature of appearance concerns in these conditions, where individuals typically focus on particular perceived feature-based flaws rather than global appearance characteristics, such as the size of the face. Additionally, facial adiposity may not evoke the same intense cognitive-affective reactions unless it aligns with the specific flaws (e.g., muscular-related flaws) that the individual is fixated on. Supporting this interpretation, Lei et al. (2019) found that muscle mass, rather than fat mass, strongly influenced perceptions of male facial masculinity in male faces. Additional research provides context for these findings. Malcolm et al. (2021) revealed that face shape concerns were more common among women than men with BDD. Although face shape relates more to facial morphology than facial adiposity *per se*, these findings might suggest that facial adiposity holds more relevance for women. While males demonstrate clear preferences for muscular, low-adipose body types (Ridley et al., 2022), and research has established connections between facial morphology and BMI/body composition (de Jager et al., 2018), these general appearance preferences may not directly translate to clinical facial appearance concerns.

Interestingly, preliminary analyses suggested that the relationship between self-face recognition and evaluations was not moderated by BDD or MD symptom severity. This suggests that recognition difficulties and negative self-evaluations may operate independently, challenging the hypothesis that more severe symptoms would strengthen the link between recognition deficits and negative evaluations. The lack of moderation could be due to several factors, including the complex and multidimensional nature of BDD and MD symptoms in ways not fully captured by our measures.

### Strengths, Limitations, and Future Directions

Methodological strengths include a sufficiently powered sample for detecting main effects, which enhances the validity of our primary findings, as well as controlling for potential confounds and incorporating both objective and subjective assessments of recognition accuracy to provide a comprehensive investigation. However, several limitations warrant consideration. First, the cross-sectional design constrains causal inferences, suggesting the need for longitudinal studies using advanced statistical approaches (e.g., network analysis) to elucidate potential mediating mechanisms.

Second, as the sample size was underpowered for detecting interaction effects in moderation analyses, these analyses should be interpreted as preliminary and exploratory, underscoring the need for future research with larger, adequately powered samples to confirm and extend these findings.

Third, the virtual administration of the SFRT potentially introduced variability in viewing conditions (e.g., screen size, lighting), possibly contributing to participants requiring more self-face information for self-recognition compared to previous in-person studies (54% vs 37-47%; Panagiotopoulo et al., 2017; Tsakiris, 2008). The SFRT’s sensitivity as a measure of objective recognition accuracy requires further validation, particularly its ability to capture subtle individual differences in self-face processing across contexts. For instance, preliminary evidence using a similar SFRT suggests that increased attractiveness ratings of an unfamiliar face lead to blurring of self–other boundaries (Panagiotopoulou et al., 2022). Additionally, our single-item, unvalidated measures of self-reported facial attractiveness and adiposity limits interpretative depth. Future research should incorporate validated tools and compare self-perceptions with observer ratings and objective measures (e.g., facial width□to□height ratio; de Jager et al., 2018) to better capture distortions in facial adiposity and attractiveness evaluation.

Fourth, generalizability is limited by our community-based sample of predominantly young, university-educated Australian males of White and East/Southeast Asian ethnicity, with a relatively low prevalence of BDD and MD risk (15-17%). Future research should examine clinical populations with verified diagnoses and more diverse socio-demographic groups, particularly considering cross-cultural variations in men’s physical appearance ideals (Mellor et al., 2014; Thornborrow et al., 2020).

Finally, future studies should explore potential underlying mechanisms, including specific mirror-checking techniques, feature-based processing biases, impaired holistic processing (Virgili et al., 2024), face-related anxiety (Bohon et al., 2012), and memory biases (Möllmann et al., 2024b). Advanced methodologies such as eye-tracking may reveal attentional biases, while experimental manipulations such as altering SFRT parameters to emphasise holistic processing could provide deeper insights into underlying mechanisms.

### Theoretical and Clinical Implications

Pending replication and extension, our findings contribute to theories of male appearance concerns through predictive coding theory (Apps & Tsarkiris, 2014). This theory suggests that self-face perception relies on predictive models integrating sensory inputs and internal representations. In BDD, self-face recognition accuracy and evaluation may be influenced by multisensory integration, where negative appearance beliefs override sensory data. This aligns with multisensory integration research showing heightened susceptibility to body illusions in those with dysmorphic concerns (Kaplan et al., 2014). This framework offers a theoretical foundation for understanding perceptual and evaluative disturbances in BDD.

Current BDD interventions primarily address distress and anxiety but neglect the broader spectrum of self-face evaluations. Our findings, linking BDD symptoms to self-reported recognition difficulties without perceptual recognition deficits, align with documented dysfunctional metacognition (i.e., awareness and regulation of one’s own cognitive processes; Wells, 2011) related to physical appearance in BDD (Zeinodini et al., 2015; Veale et al., 2014). This highlights the need for interventions to consider integrating feedback on recognition performance and metacognitive awareness training (Zeinodini et al., 2015).

The finding that appearance intolerance was associated with self-face perception suggests the need for broader conceptualizations and treatments for MD, which often focus on muscularity concerns (Cunningham et al., 2017). However, future research should clarify the role and clinical significance of self-face perception in MD.

## Conclusion

Our study reveals that self-face perception in males with BDD and MD symptoms is complex and multifaceted. Higher BDD symptoms and MD appearance intolerance symptoms were linked to greater self-reported recognition difficulties and negative self-face evaluations, while objective recognition and adiposity estimations remained intact. These findings underscore the intricate nature of self-face perception, intertwined with cognitive and emotional mechanisms. Preliminary findings also suggest that accuracy and evaluation are independent processes, operating similarly across the symptom continuum, however, cautious interpretation is warranted. Future research should explore the underlying mechanisms driving these associations, such as multisensory integration, visual processing biases, anxiety, and memory. Investigating these processes in larger and more diverse clinical samples will be essential to inform prevention and intervention strategies for BDD and MD.

## Data Availability

All data produced in the present study are available upon reasonable request to the authors

